# Comparative study between first and second wave of COVID-19 deaths in India - a single center study

**DOI:** 10.1101/2022.05.09.22274860

**Authors:** Prakash Tendulkar, Pragya Pandey, Prasan Kumar Panda, Ajeet Singh Bhadoria, Poorvi Kulshreshtha, Mayank Mishra, Gaurika Saxena

## Abstract

**Background:** The severe acute respiratory syndrome coronavirus 2 (SARS-CoV-2) is continuously evolving and many mutant variants of the virus are circulating in the world. Recurrent waves of COVID-19 have caused enormous mortality all over the world. It is of utmost importance for a health expert to understand the demographic and clinical attributes between the first and second waves of COVID-19 induced deaths.

**Method:** This was a hospital record based comparative study of baseline demographic, clinical and laboratory parameters of the first and second wave of COVID-19 in a tertiary care hospital in Uttarakhand, India. The study included all deceased patients admitted to the hospital during the first and second wave of COVID-19, i.e., between March 2020 to January 2021 and between March 2021 to June 2021, respectively.

**Result:** The study showed that there were more casualties in the second wave compared to the first, 475 (19.8%) and 424 (24.1%) respectively. There was no significant difference in terms of age. A male preponderance of mortality was evident in both the waves. The median duration of hospital stay was 5 (3-10) days in the second wave, which is significantly different from the corresponding duration in first wave (p<o.ooo). The most common clinical manifestation among the deceased were dyspnoea in both the waves, followed by fever and cough, the difference was statistically significant for cough (p< 0.000) The most prevalent comorbidity was diabetes mellitus (DM), followed by hypertension (HTN), with significant difference for HTN (p<0.003). The most frequently deranged lab parameter was lymphopenia with a significant difference across both the waves (p<0.000).

**Conclusion:** In both the first and second COVID-19 waves, older males (>45 years) with comorbidities like HTN and DM were most susceptible for COVID-19 related mortality. The study also demonstrated that most of the baseline demographic and clinical characteristics which are attributed to the mortality were more common during the second wave of COVID-19.

## Introduction

The emergence and rapid spread of coronavirus disease 2019 (COVID-19) has caused enormous mortality worldwide. This pandemic witnessed massive deaths, which the world has not seen in the last 100 years.

World health organization (WHO) declared COVID-19 a global pandemic on March 11, 2020. Since then, India has witnessed three waves. Compared to the first wave, the second wave has had severe consequences worldwide in terms of cases and mortality. Even in India, mortality was higher in the second wave than in the first wave (1). As of April 17, 2022, More than 43 million RT-PCR confirmed cases had been diagnosed in India, with more than 5.21 lakh deaths attributed to COVID-19 infection (2).

As illustrated in fig-1, the first wave had a prolonged course that was from March 2020 to approximately January 2021, whereas the second wave had a rapid course which started in the month of March-April 2021, and over the next 1-2 months, there was a sharp rise of cases and related deaths (3).

**Fig-1:**
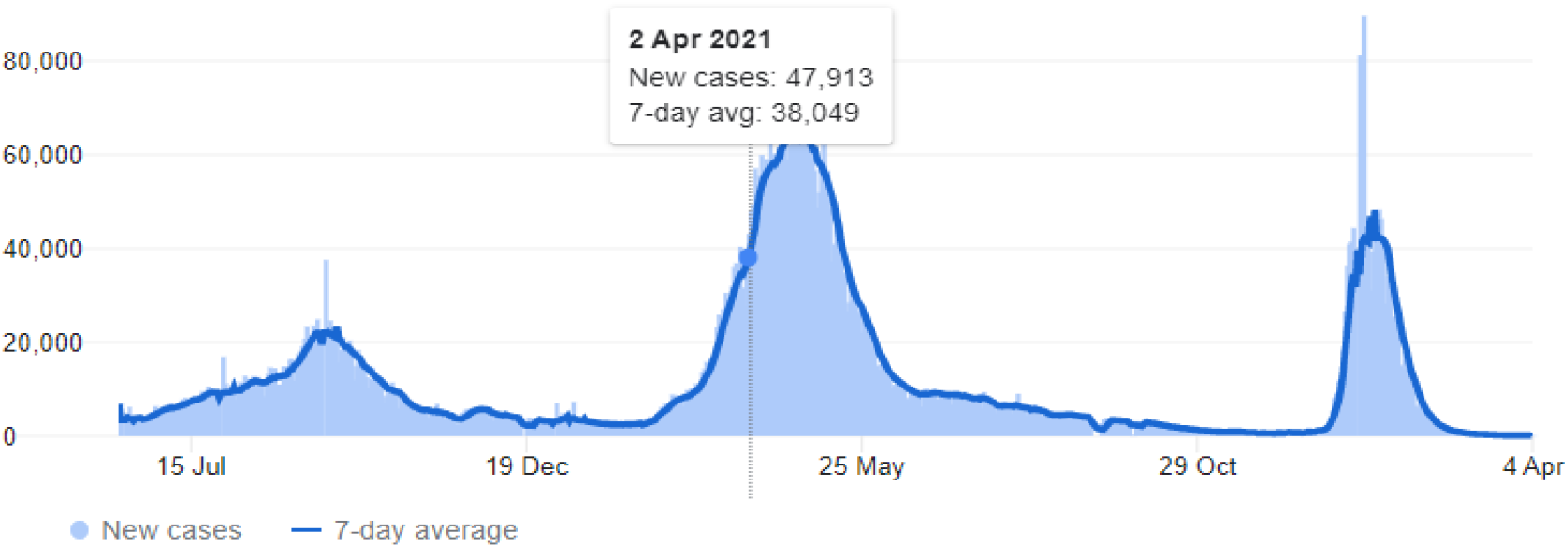
Trend of COVID-19 cases in India.

A descriptive study is already done at the same center to assess the comorbidities, risk factors, clinical signs and symptoms, and other baseline characteristics in the first wave (4). Present study is aimed to look for the difference in the baseline characteristics in first and second waves at a tertiary care hospital in Uttarakhand, India.

## Material and methods

This was a hospital record based comparative study of baseline demographic, clinical and laboratory parameters of the first (March 2020 to January 2021) and second (March 2021 to June 2021) wave of COVID-19. The study had included all deceased patients who tested positive for SARS-CoV2 by reverse transcription-polymerase reaction (RT-PCR) during the above time periods. The data was accessed from e-Hospital portal, Health Management Information System (HMIS) authorized by the government of India.

Appropriate ethical approval was obtained from the institute’s ethical committee before accessing the desired data [CTRI/2020/08/027169]. Microsoft Excel spreadsheet was used for data management. Data analysis was conducted on SPSS Statistics for Windows, Version 25.0, two proportion chi square test was performed for inferential statistics.

## Results

Out of the total reported COVID-19 cases till January 2021 (first wave), 2396 got admitted at this tertiary care hospital and 424 causalities reported. During the second wave of COVID-19, a total of 1758 patients got admitted, out of which 475 casualties were reported (Fig-2A).

**Fig-2:**
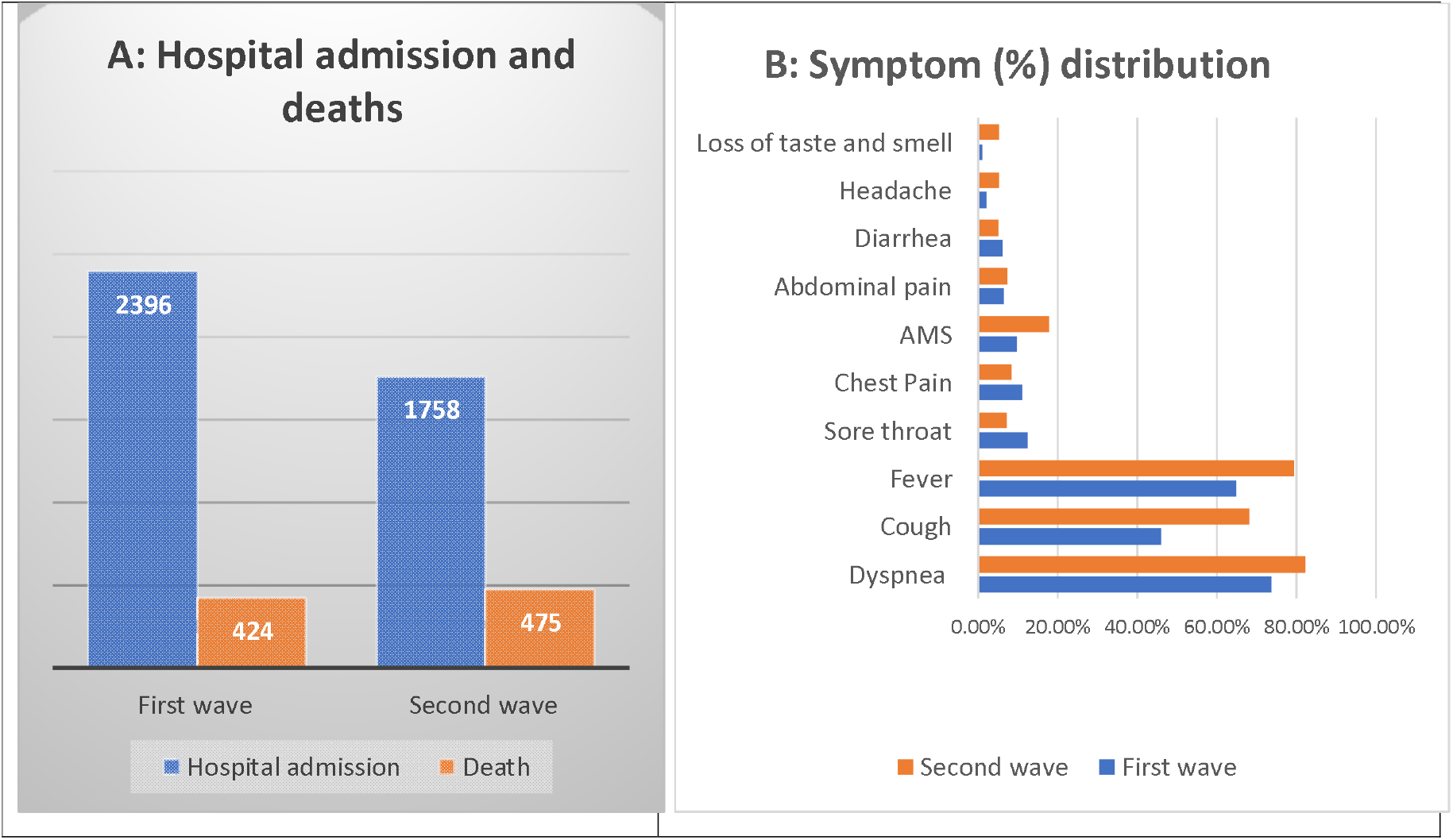
Baseline characteristics of COVID-19 deaths during first and second waves with total admission and death (A) and symptom distribution (B).

The baseline characteristics of the patients are shown in table-1. In the first wave, the mean age of the patients was 55 ± 16.24 years (95 % CI, 54.35 – 57.45), while in the second wave, the mean age in years was 56.81 ± 14.92 years (95 % CI, 55.46 – 58.15). The mean age was not statistically different among both the groups (p=0.384) Majority of deceased in both the waves were more than 45 years of age with no statistical difference among the young, middle and elderly age groups. There was significant difference in both the waves with respect to gender (p <0.004). Male preponderance of mortality was evident in both the waves. The median number of days between onset of symptom and hospital admission was 6 (3-11) days in second wave, which is significantly different from corresponding interval in first wave (p<0.000). The median duration of hospital stay was 5 (3-10) days in second wave, which is significantly different from corresponding duration in first wave (p<0.000).

**Table-1:** Baseline characteristics of patients in first and second COVID-19 waves.

|  |  |  |  |
| --- | --- | --- | --- |
| Variable | First wave<br>(N-424) | Second wave<br>(N-475) | p-Value |
| **Gender** |  |  |  |
| Male | 70.3% (65.68-74.60) | 61.3% (56.72-65.67) | 0.004 |
| Female | 29.7% (25.40-34.32) | 38.7% (34.33 - 43.28) |  |
| **Age** |  |  |  |
| <45 years | 20.51% (16.77-24.68) | 21.1% (17.47-25.00) | 0.809 |
| 45-60 years | 35.37% (30.82-40.14) | 35.2 % (30.86- 39.64) |  |
| >60years | 44.10% (39.32-48.98) | 43.6% (39.07- 48.17) |  |
| **Symptoms** |  |  |  |
| Dyspnea | 73.6%, (69.1–77.7) | 82.3% (78.58-85.64) | 0.074 |
| Cough | 46.1%, (41.1–50.8) | 68.2% (63.81-72.38) | 0.000 |
| Fever | 64.92%, (60.1–69.4) | 79.4% (75.45-82.92) | 0.228 |
| Sore throat | 12.5%, (9.98–16.43) | 7.2% (5.01 – 9.86) | 0.002 |
| Chest Pain | 11.2%, (8.51–14.59) | 8.4% (6.08-11.29) | 0.183 |
| AMS | 9.8%, (7.4–13.28) | 17.9% (14.55-21.64) | 0.002 |
| Abdominal pain | 6.5%, (4.2–8.9) | 7.4% (5.19-10.10) | 0.668 |
| Diarrhea | 6.3%, (4.04–8.69) | 5.1% (3.26-7.42) | 0.311 |
| Headache | 2.2%, (0.09–3.80) | 5.3% (5.26-7.67) | 0.025 |
| Loss of taste and smell | 1.1%, (0.15–2.20) | 5.3% (5.26-7.67) | 0.001 |
| **Sign** |  |  |  |
| Tachypnea | 80%, (76.14– 83.76) | 94.9% (92.58-96.74) | 0.000 |
| Hypoxia | 64.86%, (60.10–69.40) | 71.10% (66.86-75.19) | 0.061 |
| Hypotension | 8.1%, (5.83–11.14) | 9.47% (6.99-12.47) |  |
| **Comorbidity** |  |  |  |
| DM | 41.4% (41.4–51.1) | 42.5% (38.03-47.11) | 0.745 |
| HTN | 39.8% (35.1–44.6) | 31.6% (27.42-35.97) | 0.003 |
| CAD | 15.2% (11.8–18.8) | 10.7% (8.10-13.87) | 0.014 |
| COPD | 8.3% (6.04–11.41) | 6.9% (4.83-9.62) | 0.310 |
| CKD | 7.6% (5.43–10.60) | 8.4% (6.08-11.29) | 0.843 |
| CLD | 3.4%, (1.60–5.00) | 5.5% (3.61-7.92) | 0.118 |
| Malignancy | 5.4%, (3.26–7.58) | 3.4% (1.94- 5.41) | 0.127 |
| CND | 3.6%, (1.77–5.29) | 5.9% (3.61-7.92) | 0.102 |
| **Lab Parameters** |  |  |  |
| Lymphopenia | 90.2% (86.8–92.7) | 86.1% (82.66-89.09) | 0.000 |
| Raised ALT | 59.7%, (55.00–64.33) | 62.3% (57.79-66.69) | 0.468 |
| Raised AST | 35.1% (54.8–64.3) | 44.8% (40.31-49.44) | 0.004 |
| Leukocytosis | 50.5% (45.71–55.23) | 60.4% (55.86-64.85) | 0.008 |
| AKI | 37.7% (33.1–42.5) | 41.5% (37.00-46.05) | 0.266 |
| Anemia | 28.8% (24.46–33.08) | 28.2% (24.21-32.49) | 0.799 |
| Thrombocytopenia | 25.1% (22.21–30.61) | 35.2% (30.86-39.64) | 0.004 |
| **Hospital course** |  |  |  |
| ICU stay | 78.3%(74.0782.13) | 94.5% (92.08-96.39) | 0.000 |
| NIV Need | 55.9(51.02-60.68) | 85.5% (81.98-88.52) | 0.000 |
| Inotropes support | 54.7% (49.8–59.5) | 62.9% (58.43-67.30) | 0.010 |
| Renal replacement therapy | 10.3% (7.44-13.42) | 4.4% (2.76-6.68) | 0.000 |
| **Complications** |  |  |  |
| ARDS | 68.87%, (64.22-73.25) | 87.15% (83.81-90.03) | 0.000 |
| Sepsis | 40.33%, (35.7–45.0) | 65.05% (60.58-69.34) | 0.000 |

The most common clinical manifestation among the deceased were dyspnoea in both the waves, followed by fever and cough. Other less common symptoms were sore throat, chest pain, altered mental status (AMS), abdominal pain, diarrhoea, headache and loss of taste and smell. Symptoms in the form of cough, sore throat, AMS and loss of taste and smell were more common among deceased in second wave. The difference was statistically significant for cough (p< 0.000), sore throat (p<0.002), AMS (p< 0.002), headache (p< 0.025) and loss of taste and smell (p< 0.001) among the two groups (Fig-2B).

The most prevalent comorbidity among the deceased in both the waves was diabetes mellitus (DM), followed by hypertension (HTN). Other less common comorbidities were coronary artery disease (CAD), chronic obstructive airway disease (COPD), chronic kidney disease (CKD), chronic liver disease (CLD), Chronic neurological disease (CND) and solid and haematological malignancies. There was significant difference in both the waves with respect to HTN (p< 0.003) and CAD (p< 0.014), both of the medical comorbidities were more prevalent in the first wave (Fig-3A).

**Fig-3:**
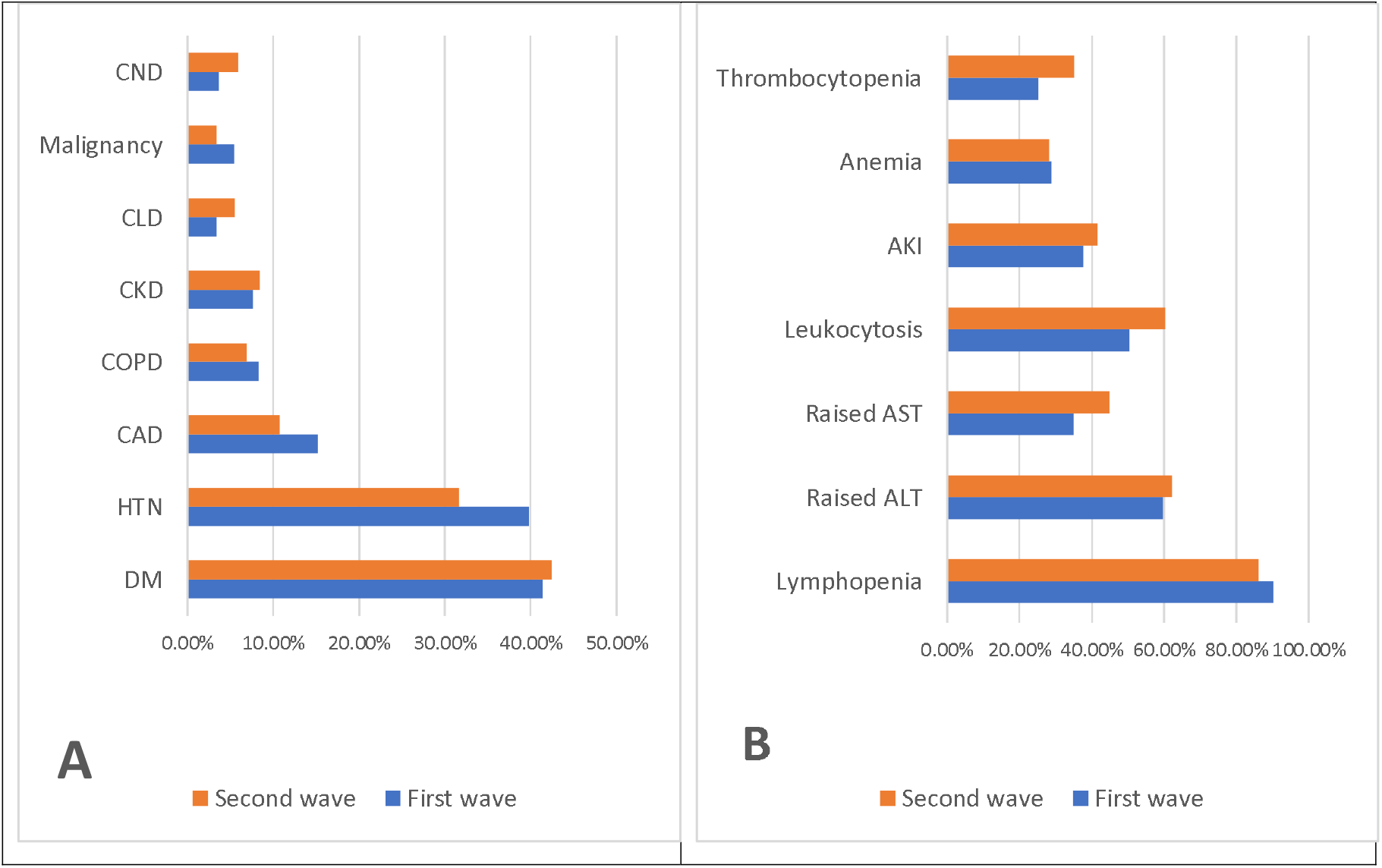
Baseline characteristics of COVID-19 deaths during first and second waves with co-morbidities (A) and deranged lab parameters (B).

The most frequently deranged lab parameter was lymphopenia among deceased in first as well as second wave. Other derangements in lab investigations were raised alanine aminotransferase (ALT), aspartate aminotransferase (AST) and leucocytosis. Acute kidney injury, anaemia and thrombocytopenia were also widespread findings among the deceased patients. There was significant difference in both the waves with respect to lymphopenia (p<0.000), Thrombocytopenia (p<0.004), raised AST (p<0.004) and leucocytosis (p< 0.008) (Fig-3B).

In-hospital course, most patients in the second wave needed Intensive Care Unit (ICU), non-invasive ventilation (NIV) and inotropic support. Complications like acute respiratory distress syndrome (ARDS) and sepsis were common during the second wave. There was statistical difference in ARDS among both the waves (p< 0.000).

## Discussion

India had witnessed the first wave of COVID-19 in 2020, which peaked in September 2020, and gradually the cases declined. Again, in March 2021, new infections with the new virus variant began to tick up and over the next 3-4 months, the country had seen chaos in the country, falling in health infrastructure and deaths in large number. According to experts, the official death toll is even higher than what the official data suggest (5). We had done this retrospective study to compare some of the baseline characteristics between the first and second wave of COVID-19.

In this study, the mean age of the patients in the first wave and the second was very similar; in the first wave, the mean age was 55 ± 16.24 years (95 % CI, 54.35 – 57.45), while in the second wave it was 56.81 ± 14.92 years (95 % CI, 55.46 – 58.15). A similar mean age group was seen in previous studies and systemic reviews; in one of the systemic reviews, consisting of 32 studies, the mean age was 56 [95% CI (48.5-59.8) years (6). In a similar review on critical patients, the mean duration of ICU stays of critical patients was 9.0 [95% CI (6.5-11.2) days (6). In one of the studies done in Pune, India, death from the diagnosis was six days (IQR, 2-11) (7). In this study, also we had a similar duration of hospital stay in both the waves. The median duration of hospital stay in the second wave was five days (IQR, 3-10), while in the first wave, it was nine days (IQR, 4-14).

Most of the previous studies have shown the male preponderance in the mortality due to COVID-19 (8,9). In our study, male preponderance was very obvious in both the waves, which was statistically significant. Even one of the systemic reviews, which included 32 studies, has shown that mortality among male patients was significantly higher with a pooled odds ratio (OR) and Hazard ratio of (HR) of 1.49 [95% CI (1.41-1.51)] and 1.24 [95% CI (1.07-1.41) respectively (10).

Older age is an independent factor for increased mortality among the patients, and several previous studies among the ICU patients have shown a similar trend (9,8). Previous research also suggests an age-dependent defect in T-cell and B-cell functions; also, there is excess production of type 2 cytokines that can lead to prolonged proinflammatory response and lead to insufficiency in preventing viral replication leading to poor outcome (11). In this study, most of the patients were more than 45 years of age. In the first wave, 79.5% of patients and in the second wave, 78.8% of patients were more than 45 years of age which is a significant population.

Dyspnea has been associated with an increased risk for COVID-19 related deaths in clinical symptoms, OR 3.31 [95%CI (1.78-6.16)] (12). Even in this study, dyspnea was the most common presenting symptom among the patients in both the waves, which is consistent with the other studies on COVID-19 mortality (13). Other common Symptoms were fever, cough, Sore throat, chest pain, altered mental status (AMS), abdominal pain, diarrhoea, headache and loss of taste and smell. In one of the studies in Iran, where they compared clinical symptoms in the first and second waves, they found that symptoms like fever, diarrhoea, loss of taste and smell and abdominal pain were more common during the second wave (14). while in this study, we found that cough, AMS, headache and loss of taste and smell was more usual symptom during the second wave, which was statistically significant.

Medical comorbidity is one of the reasons for severe events and higher mortality, and it is demonstrated in multiple studies and systemic reviews. In one of the systemic reviews, the serious event is seen in persons with hypertension (HTN) [OR-2.95, 95% CI (2.21-3.94) and with diabetes mellitus (DM) [OR-3.07, 95% CI (2.02-4.66)] (15). In another systemic review also, HTN [20.7, 95% CI (13.34-28.88)] was the most prevalent comorbidity, followed by cardiovascular diseases [9.6, 95% CI (4.81-16.23)] and DM [9.55, 95% CI (5.52-17.44)] (16). Our study had diabetes as the most common comorbidity among the patients during both covid waves with no statistically significant difference. One of the meta-analyses has demonstrated increased mortality and severity of disease in COVID-19 patients with a relative risk (RR) of 2.38 [95%CI, (1.88-3.03)] (17). Hypertension and cardiovascular disease as a comorbidity was seen more during the first wave of COVID-19 compared to the second wave, which was statically significant (p< 0.05). Other usual comorbidities which were seen in both the waves were chronic obstructive airway disease (COPD), chronic kidney disease (CKD), chronic liver disease (CLD), malignancy and Chronic neurological disease (CND), mainly cardiovascular accidents (CVA).

In this study, lymphopenia was the most common lab parameter in both the waves. Lymphopenia at presentation was reported as one of the reasons for poorer prognosis in COVID-19 as per one of the Korean studies (18).

Another meta-analysis reported that leukocytosis was more prevalent in non-survivors of COVID-19 with a weighted mean difference of 3.66 [95%, CI (2.58-4.74)]. Another meta-analysis also found that patients who had thrombocytopenia, raised alanine aminotransferase (ALT), and raised creatinine were associated with higher mortality. (12) (19) In our study, lymphocytosis, thrombocytopenia and raised aspartate transaminase (AST) were more prevalent during the second wave and which was statistically significant. As per one of the meta-analyses, thrombocytopenia increases the risk of severe COVID-19 by over five folds. (20)(21) In another meta-analysis, they found that the patients who had severe anaemia had more severe disease with a weighted mean difference (WMD) -4.08 [95%, CI (5.12-3.05)]. In this study also, anaemia was present in almost 28% of the patients.

Rapid spread and progression of the second wave had caused patients to come to the hospital in more distressing conditions and needed more aggressive management and intensive care unit (ICU) care (22,23). Most of the patients during the second wave had tachypnea and hypoxia at presentation. Also, most patients needed ICU stay and NIV support during the hospital stay. In contrast to the first wave, fewer patients underwent renal replacement therapy during the second wave. This may be attributed to the rapid progression of the disease and short hospital stays (4). ARDS remains the leading cause of death in the COVID-19 patients, as seen in multiple studies and meta-analyses including this present study (12,24).

## Conclusion

The study was a single center experience but the findings correlated with the most of the systemic reviews and meta-analysis. There were very few studies across the world which compares the baseline characteristics of the patients during the first and second COVID-19 waves. Also, there were very few studies in India. The study had showed that most of the baseline demographic and clinical parameters which are attributed with the COVID-19 mortality were more common during the second wave and can be one of the possible scientific explanations for the high mortality during the second wave in India.

## Data Availability

After obtaining approval from corresponding author, de-identified data can be shared.

## Authors’ contributions

All authors made substantial contributions to conception and design, acquisition of data, or analysis and interpretation of data; took part in drafting the article or revising it critically for important intellectual content; agreed to submit to the current journal; gave final approval of the version to be published; and agree to be accountable for all aspects of the work.

## Ethics and Data sharing

The study was done after institute ethical approval and as per declaration of Helsinki. After obtaining approval from corresponding author, de-identified data can be shared.

## Acknowledgment

A gratitude to the entire COVID-19 management team during this pandemic crisis for helping in data collection.

